# Spatially Explicit Modeling of 2019-nCoV Epidemic Trend Based on Mobile Phone Data in Mainland China

**DOI:** 10.1101/2020.02.09.20021360

**Authors:** Xiaolin Zhu, Aiyin Zhang, Shuai Xu, Pengfei Jia, Xiaoyue Tan, Jiaqi Tian, Tao Wei, Zhenxian Quan, Jiali Yu

**Affiliations:** Department of Land Surveying and Geo-Informatics, The Hong Kong Polytechnic University, Hong Kong, China; China Academy of Urban Planning and Design, Beijing, 100044, China; School of Psychology, Shenzhen University, Shenzhen, 518060, China; Beijing Engineering Research Center for Global Land Remote Sensing Products, Institute of Remote Sensing Science and Engineering, Faculty of Geographical Science, Beijing Normal University, Beijing 100875, China

**Keywords:** epidemiology, novel coronavirus, 2019-nCoV, epidemic model, disease control

## Abstract

As of February 11, 2020, all prefecture-level cities in mainland China have reported confirmed cases of 2019 novel coronavirus (2019-nCoV), but the city-level epidemical dynamics is unknown. The aim of this study is to model the current dynamics of 2019-nCoV at city level and predict the trend in the next 30 days under three possible scenarios in mainland China. We developed a spatially explicit epidemic model to consider the unique characteristics of the virus transmission in individual cities. Our model considered that the rate of virus transmission among local residents is different from those with Wuhan travel history due to the self-isolation policy. We introduced a decay rate to quantify the effort of each city to gradually control the disease spreading. We used mobile phone data to obtain the number of individuals in each city who have travel history to Wuhan. This city-level model was trained using confirmed cases up to February 10, 2020 and validated by new confirmed cases on February 11, 2020. We used the trained model to predict the future dynamics up to March 12, 2020 under different scenarios: the current trend maintained, control efforts expanded, and person-to-person contact increased due to work resuming. We estimated that the total infections in mainland China would be 72172, 54348, and 149774 by March 12, 2020 under each scenario respectively. Under the current trend, all cities will show the peak point of daily new infections by February 21. This date can be advanced to February 14 with control efforts expanded or postponed to February 26 under pressure of work resuming. Except Wuhan that cannot eliminate the disease by March 12, our model predicts that 95.4%, 100%, and 75.7% cities will have no new infections by the end of February under three scenarios. The spatial pattern of our prediction could help the government allocate resources to cities that have a more serious epidemic in the next 30 days.

## 1. Introduction

Wuhan, a large city with 14 million residents and a major air and train transportation hub of central China, identified a cluster of unexplained cases of pneumonia on December 29, 2019 ^1^. Four patients were initially reported and all these initial cases were linked to the Huanan Seafood Wholesale Market ^2^. Chinese health authorities and scientists did immediate investigation and isolated a novel coronavirus from these patients by January 7, 2020, which is then named as 2019-nCoV by the World Health Organization ^3,4^. 2019-nCoV can cause acute respiratory diseases that progress to severe pneumonia ^5^. The infection fatality risk is around 3% estimated from the data of early outbreak ^3,6^. Information on new cases strongly indicates human-to-human spread ^1,7,8^. Infection of 2019-nCoV quickly spread to other cities in China and other countries (Figure 1). It becomes an event of global health concern ^9^. Up to February 11, 2020, according to the reports published by the Chinese Center for Disease Control and Prevention, all prefecture-level cities of mainland China have confirmed cases and the total number reaches to 42667, of whom 1016 have died and 4242 recovered; 24 oversea countries have 398 confirmed cases (1 died). Chinese government took great effort to control the spread of disease, including closing the public transportation from and to Wuhan on January 23, extending the Spring Festival holiday, postponing the school-back day, and suspending all domestic and international group tours.

**Figure 1.**
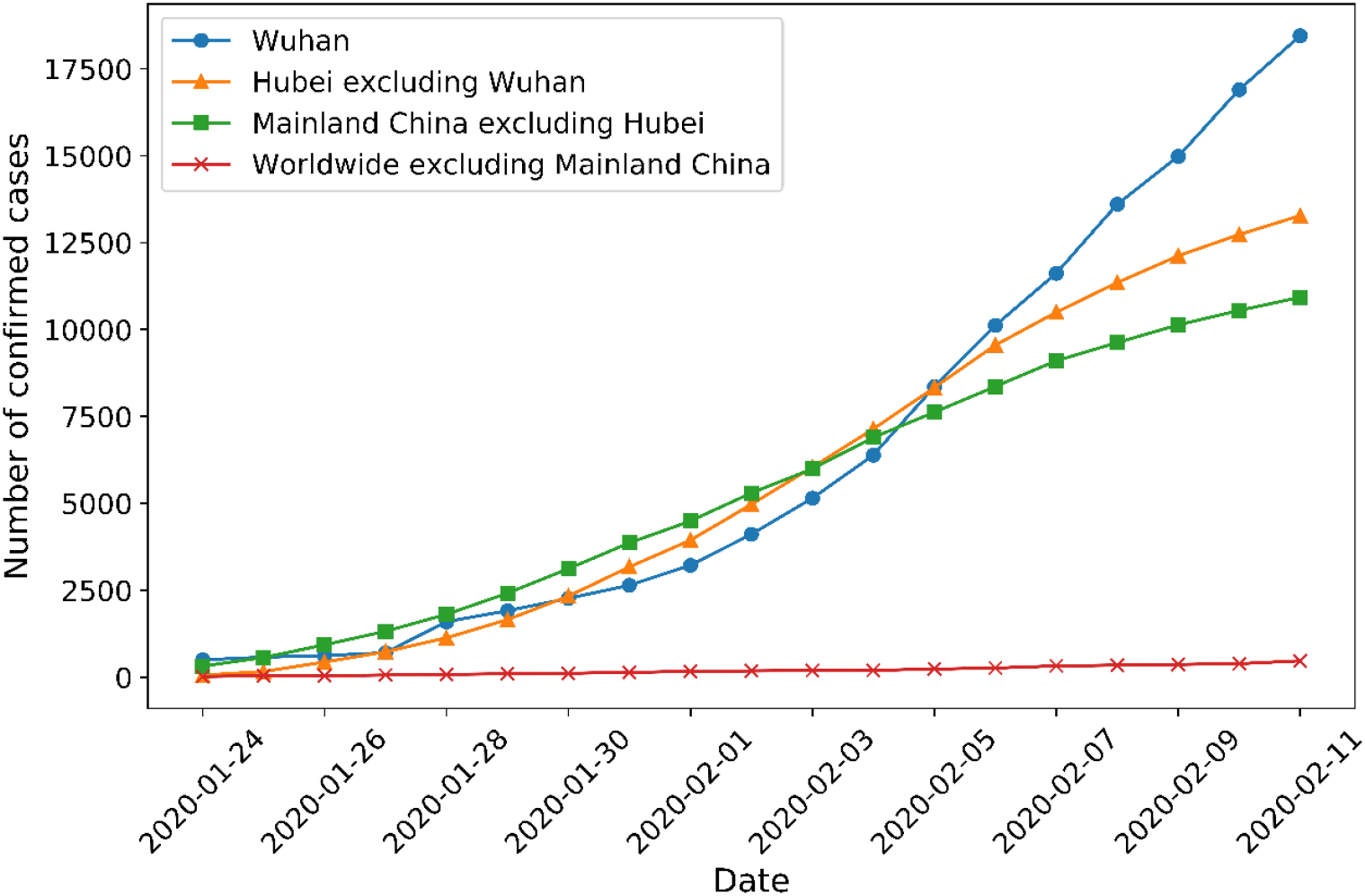
Cumulative number of confirmed cases of 2019-nCoV as of February 11, 2020, in Wuhan, Hubei province excluding Wuhan, mainland China excluding Hubei province, and outside mainland China.

Unfortunately, many external factors bring a challenge to control virus spreading. First, it might be already late to stop the migration of infected cases. Evidence suggests that Wuhan is the center of 2019-nCoV outbreak^1,10^. However, around 5 million Wuhan residents left Wuhan in January 2020 due to the Spring Festival (January 24, 2020). It is very likely that a considerable number of infected cases have moved from Wuhan to other cities before Wuhan government implemented border control on January 23. Second, it is highly possible that infected cases spread the virus to their family members or relatives ^7^. A study based on 425 patients at the early stage of outbreak revealed that the time from infection to illness onset is 5.2 days ^1^. As a result, presymptomatic cases who have left Wuhan may not be isolated themselves from their family and relatives ^11^. Third, due to the sudden outbreak of virus, the preparation and resources for preventing virus transmission are limited. The protective equipment in many hospitals in Wuhan was in short supply so that it is difficult to maintain strict personal hygiene. With the quick increase of infected cases, Wuhan and other cities in Hubei Province have large pressure to isolate and give medical treatment to infected people. All above factors make preventing the spread of 2019-nCoV even more difficult than the severe acute respiratory syndrome (SARS), another coronavirus outbreak in China 17 years ago that caused more than 8000 infections and 800 deaths.

Projecting the epidemic trend of 2019-nCoV outbreak is critical for the decision makers to allocate resources and take appropriate actions to control virus transmission. Right after the outbreak, several studies have retrieved the epidemiological parameters and predicted the future situation ^12–15^. These studies used the reported cases at the early stage of outbreak and modeled epidemic dynamics in Wuhan and nation-wide. A recent study ^10^ used air passenger data and social medium data to forecast the spread of 2019-nCoV in Wuhan and other major Chinese cities. They estimated that 75815 individuals have been infected in Greater Wuhan as of January 25, 2020 and epidemics are already growing exponentially in major cities of China with a 1-2 weeks lag time behind Wuhan outbreak. Although these studies at the early stage of outbreak help us understand the key epidemiological characteristics of 2019-nCoV, the fine-scale and updated epidemic trend in individual Chinese cities remains unknown, which is more helpful for allocating medical resources to achieve the optimal result of preventing disease spreading.

To model the updated fine-scale epidemic dynamics of all individual cities in mainland China, we proposed a spatially explicit approach. We first used mobile phone data to obtain the number of people who traveled from Wuhan to each individual city. Then we developed a new epidemiological model based the classic Susceptible-Infectious-Recovered (SIR) model to fit the dynamics of 2019-nCoV at the city level. Finally, we used this model to predict the trend under three possible scenarios: the current trend maintained, control efforts expanded, and person-to-person contact increased due to work/school resuming.

## 2. Methods

### Data sources

We collected the daily data of confirmed cases of 2019-nCoV Pneumonia in 306 prefecture-level cities in mainland China up to February 11, 2020 (Supplementary data Table 1) from a platform reporting real-time statistics of 2019-nCoV (https://ncov.dxy.cn/ncovh5/view/pneumonia). These daily reported data were used to train and validate our epidemic model. We employed China Unicom mobile phone database (https://www.cubigdata.cn) to obtain the inter-city human mobility. China Unicom is one of three largest mobile service providers in China. It has 0.32 billion users. Considering that 2019-nCoV emerged in Wuhan around January 1, 2020 and Wuhan implemented the quarantine on January 23, 2020, we collected the number of people who have Wuhan travel history during January 1-24, 2020 in each city based on the mobile phone dataset (Figure 2 and Supplementary data Table 2). In addition, Household Registered Population at 2017 year-end derived from census data was used to approximate the number of local residents in each city during 2020 Spring Festival (Supplementary data Table 2).

**Figure 2.**
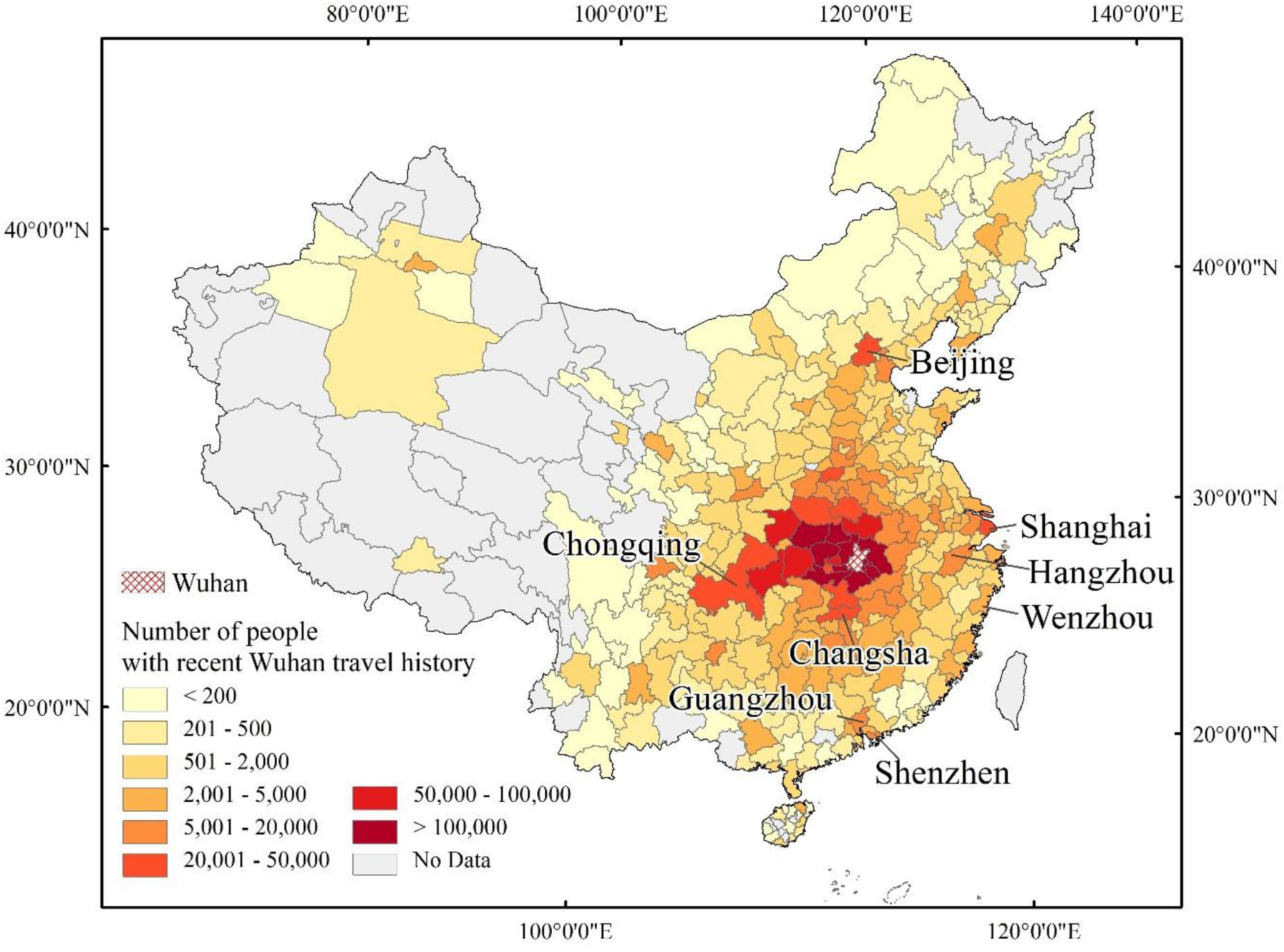
Population with Wuhan travel history in each city of mainland China during January 1-24, 2020

### A spatially explicit epidemic model

Our proposed model stems from the SIR model, a classic approach to simulate epidemiological dynamics. We modified the SIR structure based on the unique characteristics of the outbreak of 2019-nCoV. First, in all cities other than Wuhan, the initial infectious cases are most likely imported from Wuhan^10^. Second, many Wuhan residents moved to other cities due to the Spring Festival and this mobility was closed after the quarantine on January 23. Third, those people from Wuhan have low contacts with local residents because Chinese government required them to implement self-isolation. Last, during the past 40 days, all cities took efforts to control the virus spreading, which slows down the daily increase of new infections (Figure 1). Accordingly, in the modified SIR model, the susceptible variable *S* was divided into two groups: *S*_1_, the number of local susceptible, and *S*_2_, the number of susceptible with Wuhan travel history. These Wuhan-inbound groups (*S*_2_), have transmission rate (*β*_2_) different from transmission rate (*β*_1_) of local residents (*S*_1_) as Chinese government took measures to reduce the person-to-person contacts. A decay rate *a* was introduced to tune the value of *β* in each day, accounting for the gradual impact of prevention interventions in each city. In our modified SIR model, recovered population *R* was extended to include those cured, died, and isolated in hospital because they cannot transmit the virus. The differentiate equations of our modified SIR model is as follows:

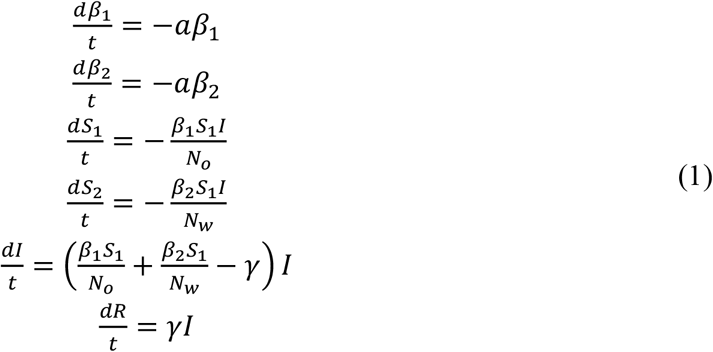

Where *N*_*o*_ is the total local population derived from the census data, and *N*_*w*_ represents the total population with Wuhan travel history during January 1 to January 24, estimated from mobile phone data; *I* is the number of infections, *β* and *γ* denote the daily transmission rate and daily recovery rate respectively.

In our model, four variables need to be initialized: (1) initial number of infectious *I*_0_, treated as a parameter to be estimated (see next section); (2) initial number of local susceptible *S*_10_, equal to the total number of the local population of each city *S*_10_ = *N*_*o*_; (3) initial number of susceptible traveling from Wuhan *S*_20_, equal to the population from Wuhan excluding the initial infectious *S*_20_ = *N*_*w*_ − *I*_0_; and (4) initial number of removed *R*_0_, assuming no cured, hospitalized, or death at initial state *R*_0_ = 0.

### Estimation of model parameters

Our modified SIR model has four parameters: transmission rate *β*_1_ among local residents, *β*_2_ among people with Wuhan travel history, decay rate *a*, and recovery rate *γ*. For *γ*, we assume that once an infected individual is hospitalized, the person will be segregated and therefore no longer infectious. According to a recent study using the first 425 patients ^1^, the mean incubation period of 2019-nCoV is 5.2 days, and the mean duration from illness onset to hospital admission is 9.1 days. We assume that the incubation period and duration from illness onset to first medical visit is similar with these 425 infected cases. Therefore, the estimated infectious period is 5.2 + 9.1 = 14.3 days and *γ* equals 1/14.3 = 0.0699.

For parameters *β*_1_, *β*_2_, and *a*, we used the daily cumulative confirmed cases up to February 10, 2020 to retrieve their optimal values to reflect the current dynamics in each city. We first estimated the optimal value of *β*_2_ and *a* of Wuhan since its epidemic model only has one transmission rate *β*_2_. Then, the estimated *β*_2_ was used as a prior parameter for the estimation of *β*_1_ and *a* for individual prefecture-level cities. The Nelder-Mead algorithm ^17^ was employed to estimate parameters through minimizing the sum of squared differences between the simulated and actual daily cumulative confirmed cases. To better capture the current trend, the number of daily cumulative cases were also used to weight the samples in parameter retrieval. Since the epidemic model is highly sensitive to the initial infectious number *I*_*0*_, and the reported initial infectious number often has large uncertainty, we treated the initial infectious number *I*_0_ as another parameter to be estimated together with *β*_1_ and *β*_2_. Specifically, we used daily cumulative confirmed cases of each city from January 25 to February 10, 2020 to retrieve the parameters and assumed January 20, 2020 as the start point when massive inter-city mobility happened before the Spring Festival. The goodness of model fitting was assessed by comparing the number from model simulation and reported cases. The trained model was further validated using the reported data on February 11, 2020.

### Prediction of different scenarios

We predicted the epidemic dynamics in the next 30 days under three scenarios: the current trend maintained (scenario 1), control efforts expanded (scenario 2), person-to-person contacts increased due to work resuming (scenario 3). These three scenarios were designed by considering the joint effect of virus transmissibility and outbreak control ^18,19^ and realized by manipulating model parameters from February 11, 2020 to March 12, 2020 (see an example in Figure 3). Decay rate *a* reveals the effectiveness of government control and removal rate *γ* represents the promptness of medical treatment. Therefore, scenario 1 keeps *a* value the same as the trained model. In scenario 2, we doubled the value of *a* to reflect more efforts in each city for controlling the disease. In scenario 3, the interference of work resuming was considered, so a short rebound was introduced to the transmission rate *β* (i.e. changing *a* to -*a* during February 11-15 period). *γ* in all scenarios gradually increased from 1/14.3 to 1/9.8 in the 30 days to reflect the reduction of average diagnostic isolation time ^20^.

**Figure 3.**
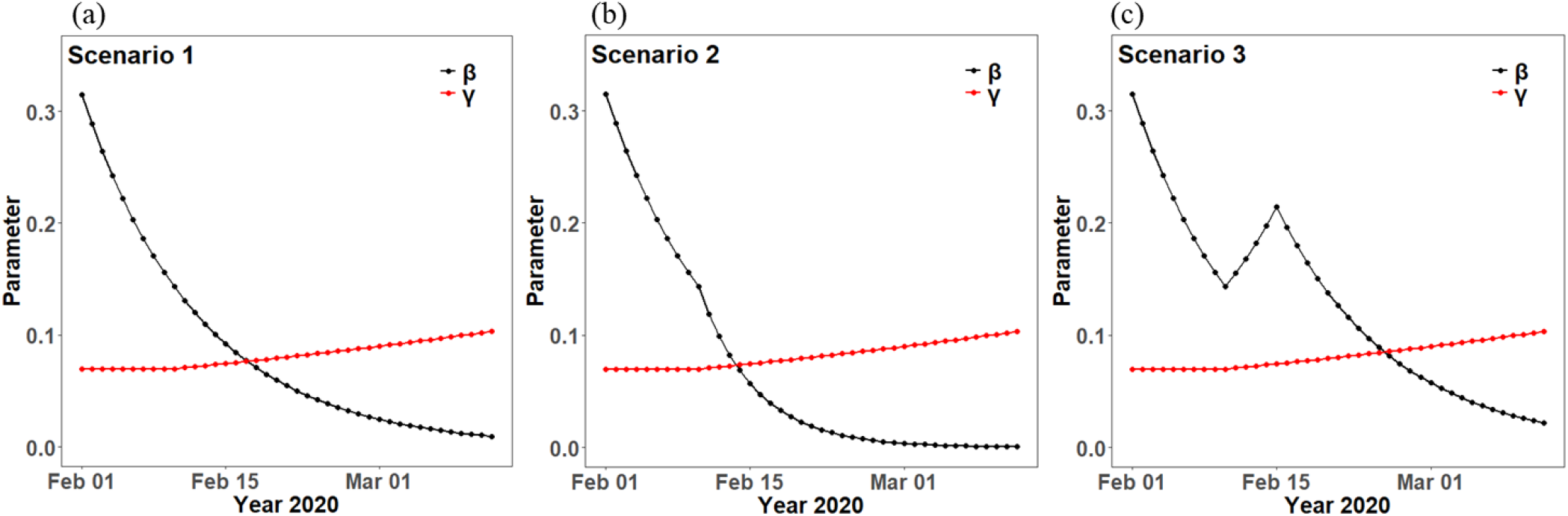
An example of temporal variation of *β* (controlled by the decay rate *a*) and *γ* under three scenarios: (a) the current trend maintained, (b) control efforts expanded, (c) person-to-person contacts increased due to work resuming

## 3. Results

### Parameter estimation

To examine the goodness of model fitting, we calculated R-squared and root mean square error (RMSE) between the fitting result and confirmed cases in each city (see representative examples in Figure 4). The median value of R-squared and RMSE across all cities is 0.96 and 1.20 respectively, indicating that our model can well fit the current spreading trend. We examined the fitted models with R-squared less than 0.7 (26 out of 306 cities) and found that all these cities have very small number (1-3) of confirmed cases and the number does not change in the past several days. It is reasonable to assume that these cities have completely controlled the spreading and no new infected cases will emerge in the future. Therefore, for these cities with R-squared less than 0.70, the *β*_1_ value of the city was set to zero.

**Figure 4.**
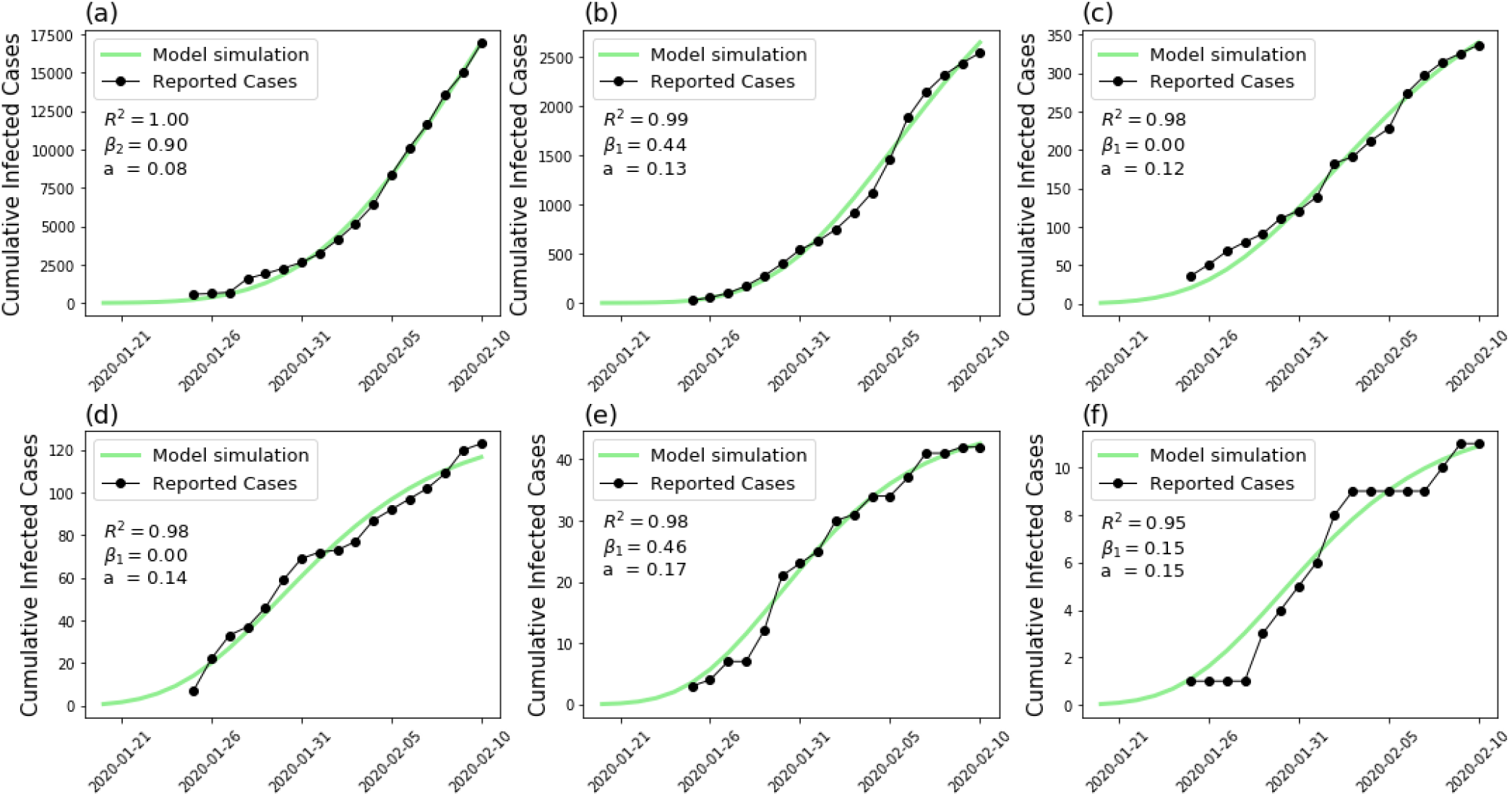
Comparison between the number from model simulation and reported cases in cities with representative severity of disease spreading: (a) Wuhan, (b) Xiaogan, (c) Beijing, (d) Chengdu, (e) Kunming, and (f) Datong

Transmission rate *β*_2_ estimated using Wuhan confirmed cases is 0.9, reflecting a high transmission around January 20. The transmission rate *β*_1_, decay rate *a*, and initial infectious population *I*_0_ vary from city to city (Supplementary data Table 3). As the transmission rate among local residents in each city, *β*_1_ reflects the intensity of control measures adopted by each local government at the beginning of outbreak, as well as the awareness of citizens to take protective measures. For example, *β*_1_ in megacities such as Beijing, Shanghai, Guangzhou and Shenzhen are low (Figure 5.a) which may attribute to higher health literacy of their citizens ^21^, although they have intensive traffic and population mobility. Decay rate *a* reflects the continuous effort input by each city to control the transmission of disease. It shows that decay rates of cities close to Wuhan is generally lower than other cities (Figure 5.b), suggesting the big challenge faced by these cities to control the disease spreading.

**Figure 5.**
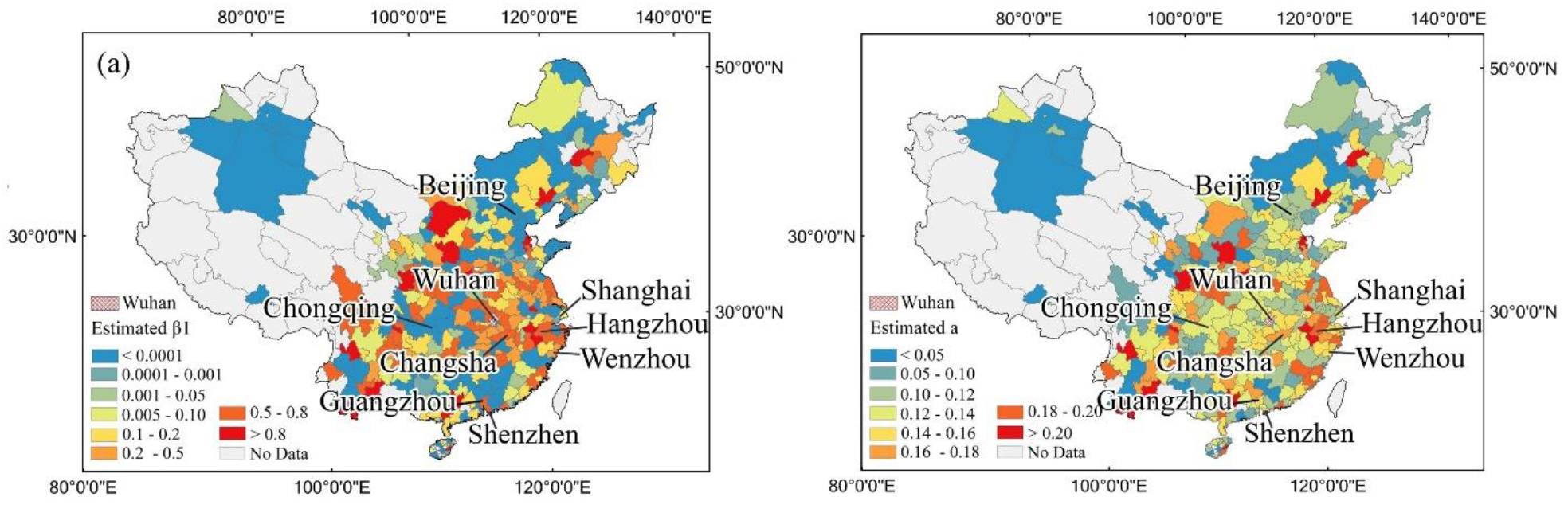
Results of parameter estimation: *β*_1_ (a) and *a* (b) of cities estimated using daily confirmed cases

### Model validation

To test the prediction capacity of our model, we used the reported confirmed cases on February 11 in all cites excluding Wuhan because its high confirmed case leads to bias in assessment. We compared the predicted daily new infections on February 11 with the reported confirmed cases in the same day. The predicted values by our model well match the reported infected cases (R-squared: 0.86, RMSE: 6.35, P < 0.0001, Figure 6.a). To demonstrate the effectiveness of our model, the results was compared with the prediction from the classic SIR model that uses uniform parameters estimated from nationwide data (*β*= 0.209, *γ*=0.0699). Compared with our model, the predicted values by the classic SIR model does not match with the daily new infected cases (R-squared: 0.30, RMSE: 13.22, P < 0.0001, Figures 6.b).

**Figure 6.**
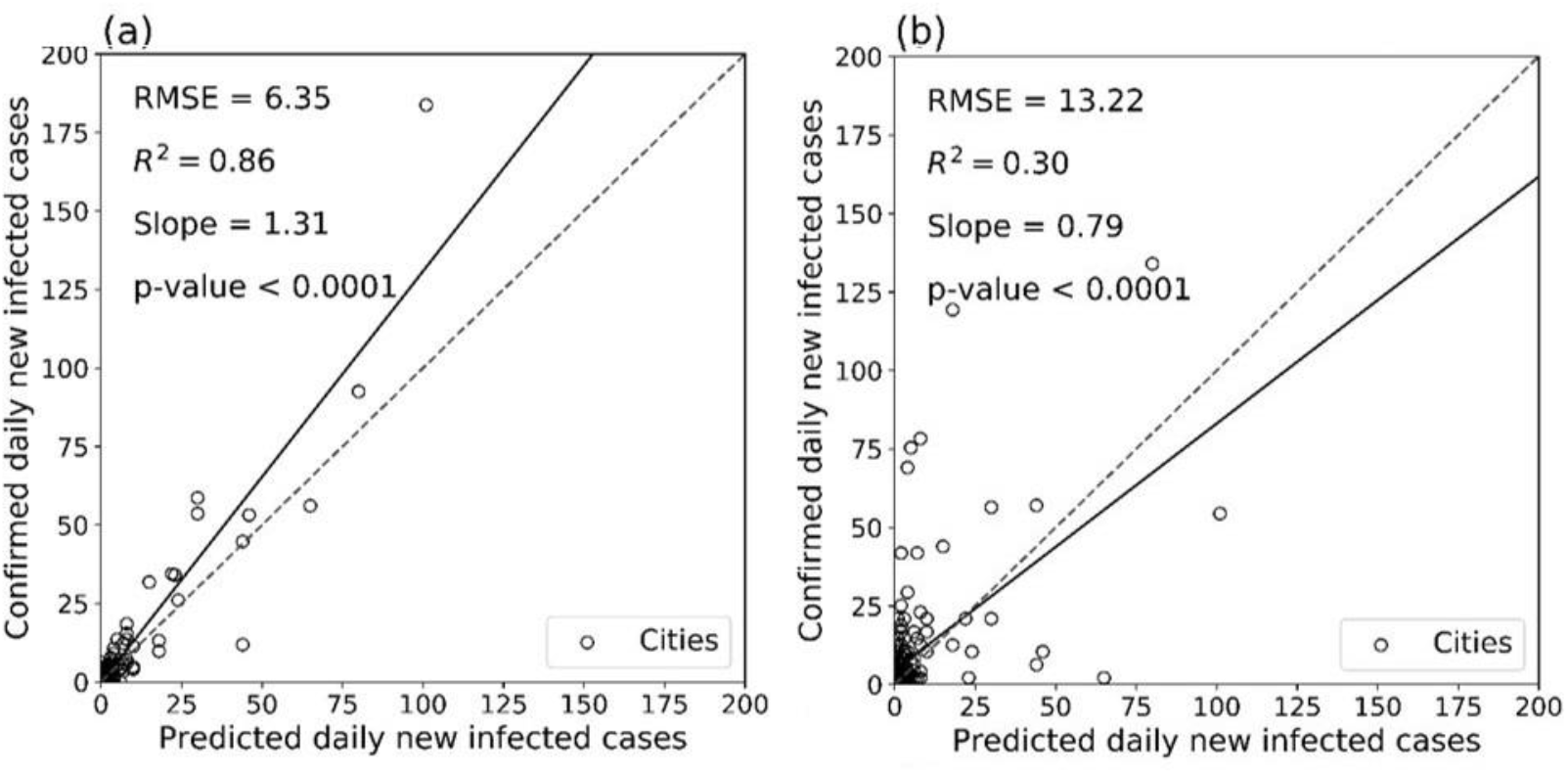
Model validation of the proposed model (a) and the classic SIR model (b) based on data on February 11, 2020. The solid lines represent the fitted linear regression line and dashed lines represent 1:1 lines for reference.

### Epidemic dynamics under three scenarios

We predicted daily infected cases of each city up to March 12, 2020 under three different scenarios (scenario 1 - the current trend maintained; scenario 2 - control efforts expanded; and scenario 3 - person-to-person contacts increased due to work resuming). Our prediction shows that the whole mainland China will have 72172, 54348, and 149774 people infected up to March 12, 2020 under the above three scenarios respectively (Supplementary data Table 4). To provide an intuitive picture about epidemic dynamics in different scenarios, we showed in Figure 7 the number of cumulative infections in each city on March 12. The infected people will mainly distribute in the central and eastern provinces, the number of western cities at a relatively low level under all scenarios. In scenario 3 (Figure 7.c), many cites have much larger number of infections than scenario 1 (Figure 7.a) and 2 (Figure 7.b), suggesting that work resuming will bring large challenge to control the disease timely. This difference is significant for those cities with largest number of infections by March 12 (Figure 8), such as Wuhan and other cities in Hubei province.

**Figure 7.**
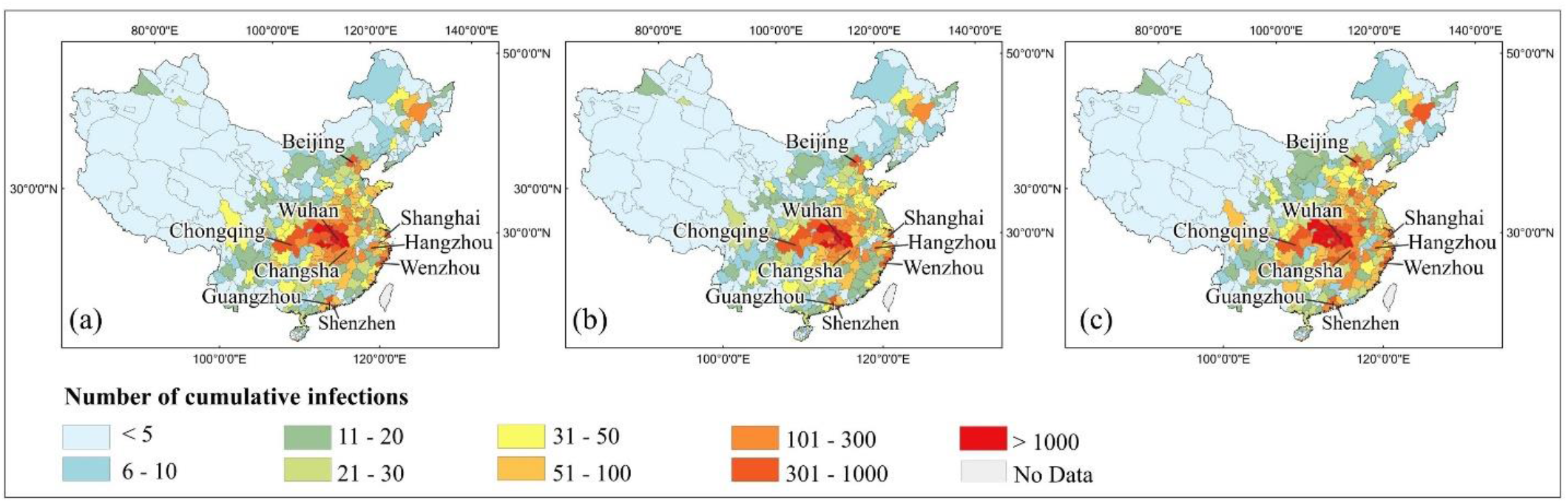
Mapping of predicted cumulative infections on March 12: (a) scenario 1, (b) scenario 2, (c) scenario 3

**Figure 8.**
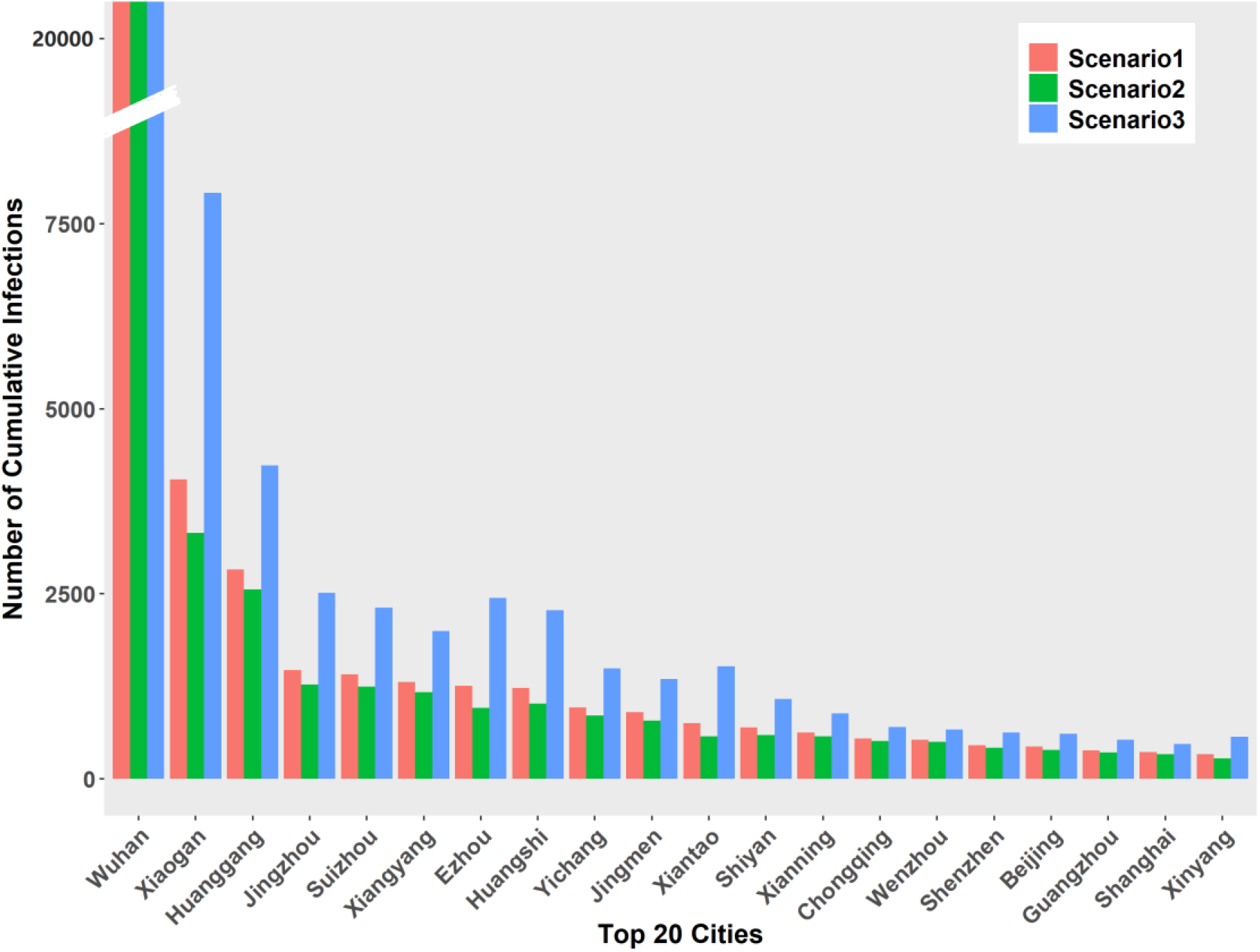
20 cities with highest cumulative infections by March 12 predicted under three scenarios (Data of Wuhan is 40613, 27021, and 98486 that is not shown properly in this figure)

To understand the specific attributes of epidemic dynamics under different scenarios, we investigated the temporal changes of daily new infections across all cities in mainland China. In Figure 9, we show the results in Wuhan, Hubei province excluding Wuhan, other provinces, and four first-tier cities. Compared with the scenario 1 where current trend is maintained, the daily new infections in scenario 2 reduces quickly in the second half of February. In scenario 3 where transmissibility rebounds after the public holiday in all cities, the peak of new infections will postpone ten days and the magnitude will be twice of that in scenarios 1 and 2. Our simulation suggests that strict quarantine of inner- and inter-city population movement during February would have a significant effect on the suppression of virus spreading.

**Figure 9.**
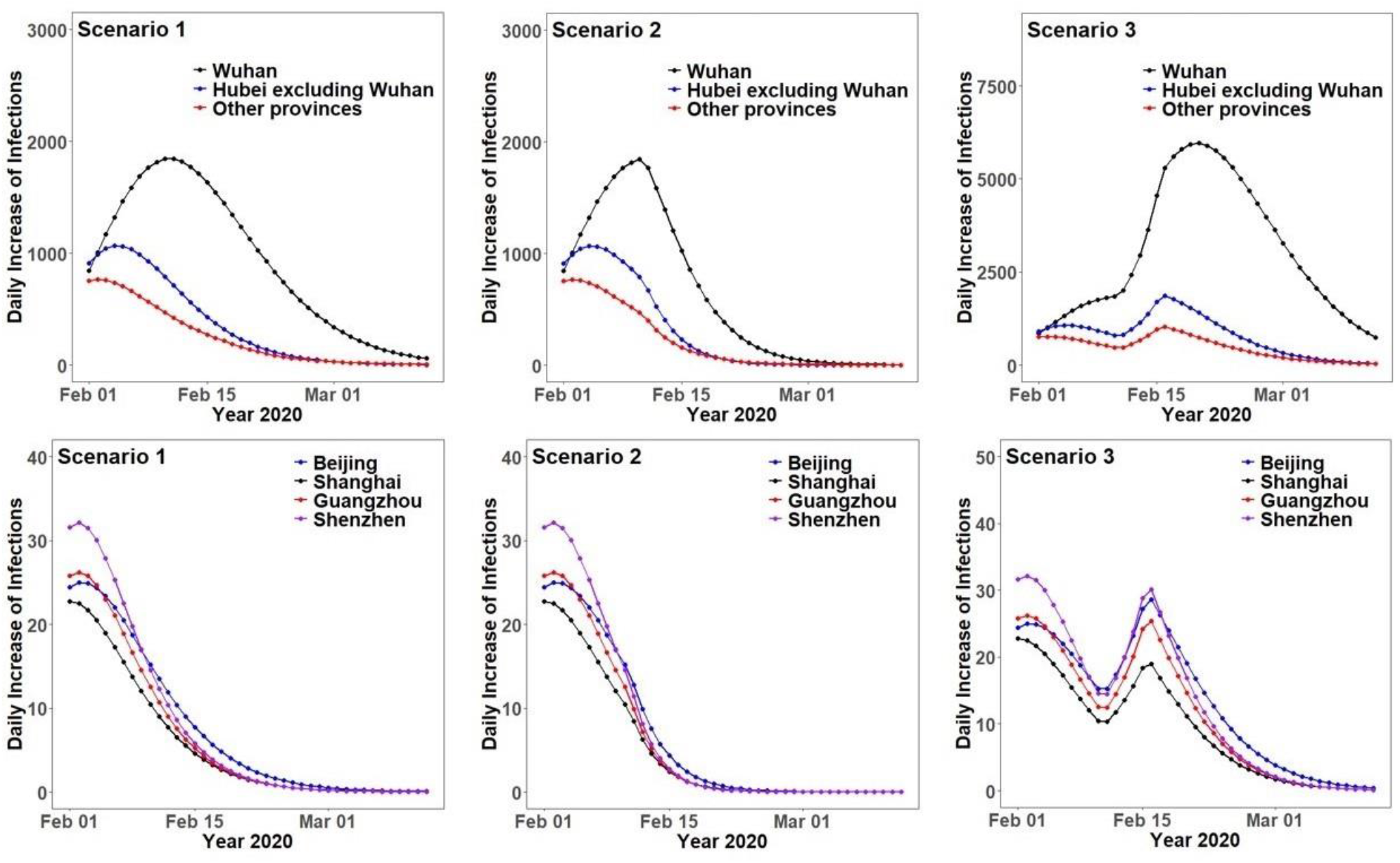
Temporal change of daily new infections in Wuhan, Hubei province excluding Wuhan, other provinces, and four first-tier cities based on the proposed model

Change of daily new infections number is an important indicator of effective intervention. The date when the number of new daily infections reaches the peak (Figure 10. a-c, Supplementary data Table 4) suggests that the exponential growing of disease will stop and the spreading would be controlled gradually. Under current trend, our model estimated that the number of new infections in 79.7% cities already reached the peak point before February 11 and in all other cities, it will reach the peak point by February 21. With the control effort expanded, all cities will have the peak point of new infections by February 14, one week earlier than current trend. However, the peak point of new infections will be greatly delayed under scenario 3 that a few cities in Hubei province will show the peak point of new infections by February 26.

**Figure 10.**
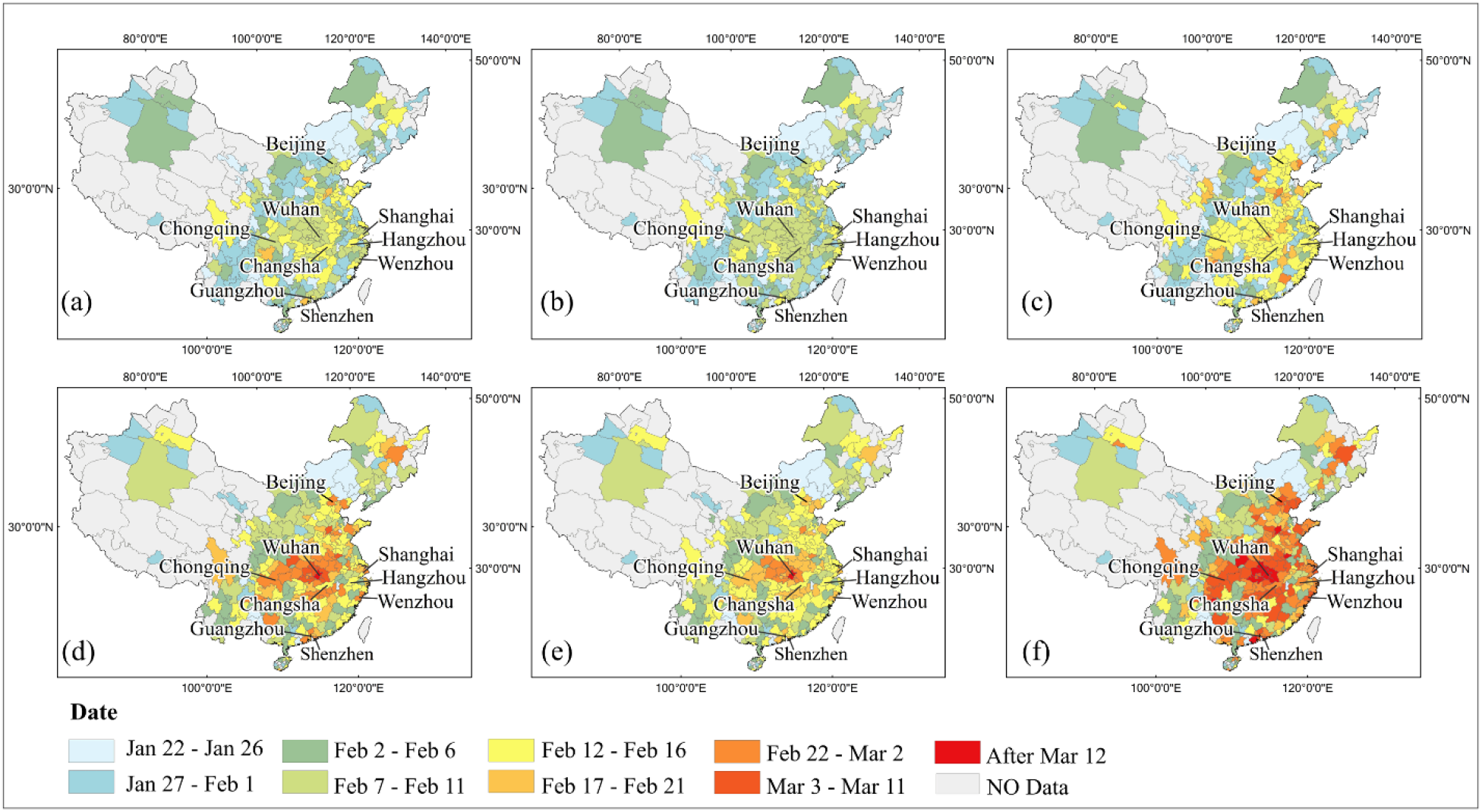
Dates when the number of daily new infections reaches the peak point under scenario 1(a), scenario 2(b), and scenario 3(c); dates when no more daily new infections emerge under scenario 1(d), scenario 2(e), and scenario 3(f).

The daily new infections decreasing to zero (Figure 10.d-f, Supplementary data Table 4) is the signal of eliminating the coronavirus outbreak. Our results show that this successful day in cities close to Wuhan is much later than other cities under all scenarios. Our prediction suggests Wuhan cannot eliminate the disease by March 12 under all scenarios. For other cities, if the current trend is maintained (scenario 1), 95.4% cities can eliminate the disease by the end of February and all of them can succeed by March 12. With control efforts expanded (scenario 2), all cities except Wuhan will completely control the disease by the end of February. However, only 75.7% cities can completely win the battle against 2019-nCoV by the end of February if person-to-person contacts increase due to work resuming (scenario 3). Nine cities in Hubei province still have daily new infections on March 12, 2020, the last day in our prediction under scenario 3.

## 4. Discussion

In this study, we modeled the updated epidemic trend of 2019-nCoV for each individual city in mainland China and used the model to predict the future trend under three possible scenarios. Our model accounts for 86% of the variation in the number of daily reported infections on February 11 across cites in mainland China. With this model, we predict that all cities will show the peak point of daily new infections by February 21, if the current trend is maintained. This date can be advanced to February 14 with control efforts expanded or postponed to February 26 under pressure of work resuming. Cities in central and east of mainland China will face a challenge to prevent the growing of disease transmission due to work resuming.

Compared with other recent modeling studies, our study has the following strength. First, the unique characteristics of the novel coronavirus spreading were taken into account in our proposed model. Considering that the new coronavirus originates from Wuhan and the majority of the local infected patients outside Wuhan had Wuhan travel history before the Spring Festival, we extended the SIR model to capture the transmission characteristics of the novel coronavirus in mainland China. Second, a decay rate was introduced into the model for quantifying the intervention implemented in each city. Third, our proposed spatially explicit model is able to obtain fine scale prediction result. Different cities have different transmission rates due to their own conditions (for instance, population density and human mobility characteristics). If only the transmission rate in national scale is used to model the epidemics of different cities ^10^, the prediction of epidemic trend of all major cities would be similar to Wuhan (see Figure 4 in Wu et al., 2020).

Our predictions of three future scenarios, namely the current trend maintained, control efforts expanded, person-to-person contacts increased due to work resuming, provide information for decision makers to allocate resources for controlling the disease spread. Generally speaking, densely populated cities and cities in central China will face severe pressure to control the epidemic, since the number of infections keeps increasing in all three scenarios in the near future. By comparing predictions of three scenarios, it is obvious that reducing the transmissibility is a critical approach to reduce the daily new infections and controlling the magnitude of epidemics. Fortunately, the latest number of confirmed diagnoses (Figure 1) and our prediction both show the slowing down of new infections in these days, indicating current control measures implemented by Chinese government are effective, including controlling traffic between Wuhan and other regions, isolating suspected patients, canceling mass gatherings, and requiring people to implement protective measures. However, once the Spring Festival travel rush returns as scenario 3 (most provinces planned to resume work on February 9), it will inevitably cause considerable growth in transmissibility and further re-increase of epidemics. In addition, current insufficient supply of protective equipment may exacerbate this situation. Therefore, public health interventions should be performed continuously to obtain the best results of epidemic control. The following measures are recommended to implement continuously in the near future, such as, postponing work resuming, arranging work-from-home, and instructing enterprises to implement epidemic prevention measures. Essentially, all measures are for reducing population mobility and person-to-person contact, and there is no panacea for all conditions, hence interventions in different regions should be adapted according to local epidemics.

Our modeling work has several limitations. First, due to the limited prior knowledge for this sudden 2019-nCoV outbreak, the infection rate and recovery rate in this study are regarded as the same for different age groups, which may result in errors of predication for cities with different age structures. Second, the model parameters were estimated using the reported confirmed cases that may be lower than the actual number of infections, so parameter estimation may not represent the real situation. Third, besides transmission between Wuhan and other cities, we do not consider other inter-city transmissions. Although the Chinese government strictly controlled the traffic between cities, the inter-city transmission may contribute to the epidemic dynamics in future days, especially during days of work resuming.

## Data Availability

We will share all data used in our study

## Author Contributions

XZ designed the experiments. PJ, ZQ, and JY collected and processed data. AZ and SX analyzed data. All authors interpreted the results and wrote the manuscript.

## Data sharing

data obtained for this study will be available to others.

